# Pre transplant malnutrition predicts post transplant respiratory complications in living donor liver transplantation for biliary atresia- single center retrospective analysis of 110 children

**DOI:** 10.1101/2020.04.14.20061630

**Authors:** BB Vasavada

## Abstract

**Back Ground:** Biliary atresia is commonly associated with malnutrition and failure to thrive. Very few studies have been published on impact of preoperative malnutrition of post transplant outcome in these children.

**Material and Methods:** 110 children underwent living donor liver transplantation between January 2003 to march 2013.Pre transplant malnutrition was defined according to z scores for weight for age and height for age as per who definition. Patients having both Z score of < −2 were compared with control group. Statistical analysis were done using SPSS version 21 (IBM).

**Results:** 39 children out of 110 were having z score for weight for age < −2.There was no statistical difference between PELD score, graft weight, GRWR, intraoperative blood loss between to groups. 22 out of 39 patients in malnourished group developed clavein grade 3,garde 4 complications and 32 patients out of 71 in control group developed clavien grade 3 grade 4 complications. (p= 0.318). Over all mortality rate was 4.5% and mortality rates in malnourished vs control group was respectively 7.69% and 2.81% (p= 0.278). Total 14 patient developed postoperative pulmonary complications. Pulmonary complications were significantly high in malnourished group. p=0.003.

**Conclusion:** Preoperative malnutrition is associated with high postoperative pulmonary complication rate in liver transplantation for biliary atresia.

## Introduction

Biliary atresia is the most common indication of liver transplantation in pediatric population(1,2). Timely intervention such as kasai hepatoportoenterostomy can halt the progression of disease, however despite this procedure progressive hepatic damage continue in majority of patients and 70-80% patients require liver transplant within first two years of life (3,4,5). Malnutrition and growth retardation are significant problems in biliary atresia patients. Decreased oral intake, early satiety due to organomegaly, fat malabsorption, and increased energy expenditure due to a hypermetabolic state all likely contribute to malnutrition in biliary atresia patients (6). However there are few studies on impact of pre transplant nutrition status on post transplant outcome. Purpose of our study was to evaluate impact of pre transplant malnutrition on post transplant outcomes of the patients.

## Material and Methods

### Study design

Data of all the patients who underwent living donor liver transplant for biliary atresia between January 2003 and April 2013 were collected and retrospectively analyzed. We included all the patients who has z scores for weight/age and height /age −2 as study group. In end stage liver disease due to ascites, edema weight for age alone may not be the accurate indicator of nutrition status. So to avoid confusion we selected patients having both the z scores = −2 as inclusion criteria. (Wasted and shunted as per who definition) Z score was calculated as per centers of disease control and prevention charts. (http://www.cdc.gov/growthcharts/zscore.htm). Liver transplant outcome was measured as clavien dindo classification grade 3 and grade 4 complications and over all mortality (7). These outcomes were compared between malnourished and control group.

### Statistical Analysis

Statistical analysis was done using chi square test for categorical variables and Mann Whitney U test for continuous variables. P value < 0.05 was considered to be statistically significant. Multivariate analysis was done using MANOVA method. Statistical analysis was done using SPSS version 21(IBM).

### Results and Statistical analysis

110 patients underwent living donor liver transplant in period of January 2003 to April 2013.39 patients out of 110 were having Z score of weight by age and weight by height - 2.characteristics of both the group is described in table 1. Patients’ characteristics is described in table 1.Malnutrition group had significant higher median PELD (p=0.001) and Large for size grafts (GRWR >2.5) were significantly more common in malnutrition group as we do not use reduced grafts.

**Table 1.** Patients characteristics.

| Factors | Malnutrition/growth Failure group (Z= -2)(N=39) | Non malnutrition Group (N=71) | P value |
| --- | --- | --- | --- |
| Age (month)<br>(Median)(Range) | 13(2-63) | 17(3- 180) | P=0.290 |
| Sex | 21 male 18 female | 34 male 37 females | P=0.456 |
| CTP | 8.5(5-15) | 8(5-17) | P=0.815 |
| PELD | 18(8-32) | 11(5-25) | P=0.001 |
| Type of graft | 38-left lateral,1=left | 59=left<br>lateral,3=left,6=right | P=0.137 |
| Large for size<br>grafts (GRWR<br>>2.5)(n) | 34/39 | 35/71 | P=0.000 |
| Intra operative<br>blood loss(ml) | 205(200-2400) | 200(100-1500) | P=0.716 |
| Primary<br>transplant<br>(without<br>previous kasai<br>procedure) | 3 | 3 | P=0.459 |
| Age at kasai<br>(month) | 60(2-120) | 60(2-140) | P=0.959 |
| Transplant after<br>second kasai<br>procedure. (N) | 5 | 5 | P=0.337 |

Table 2 mentions various complications in two groups. 22 patients suffered from 43 complications in malnutrition group where as 32 patients suffered from 60 complications in the control group. There was no statistical significant difference between over all complication rates. (P=0.318). As can be seen from table 2 respiratory complications were significantly more common in malnourished group. (p=0.003) We included ARDS and postoperative pneumonia and atelectasis in respiratory complication group.3 patients died in malnutrition group and 2 patients in no malnutrition group. There was no statistically significant difference between both the groups in overall mortality rates. (p=0.278)

**Table 2:**
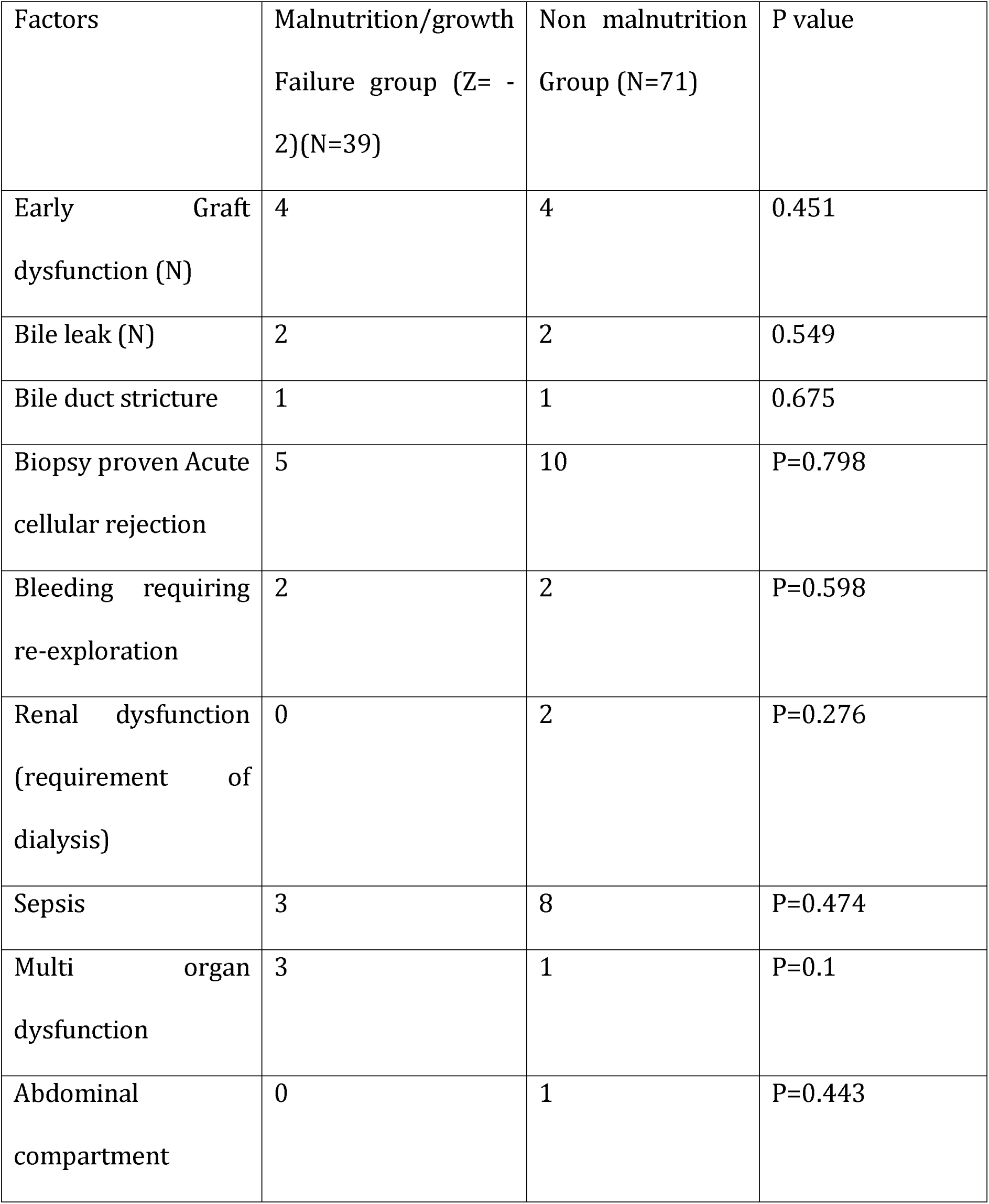
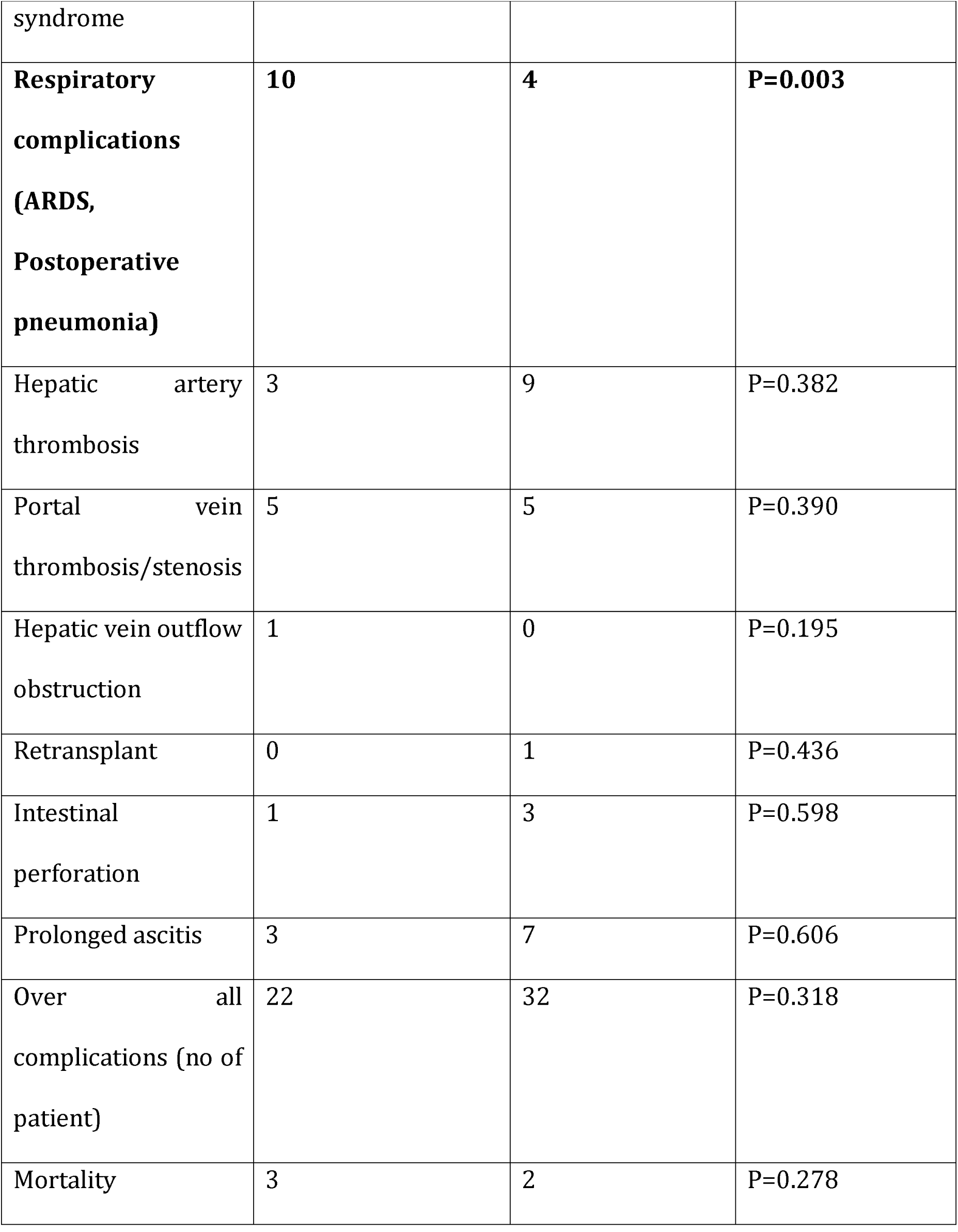
Complications observed during study period. Early graft dysfunction was defined as bilirubin greater than 20 on POD 7.

### Univariate analysis of Respiratory complications

On Univariate analysis we analyzed association between various factors like age at transplant, sex, PELD score, CTP score, Large for size graft, Operative blood loss, Operative duration, Previous kasai procedure, Timing of kasai procedure. Respiratory complications were significantly associated with PELD score (p=0.049), and Z score - 2(p=0.003). Large for size graft showed borderline P value of 0.069 and as large for size graft were significantly more common in malnutrition group we included it in multivariate logistic regression.

### Multivariate analysis: (TABLE 3)

As shown in table 3 on multivariate analysis only Z score-2 (both weight/age and height/age) independently predicted respiratory complications with p value of 0.03.

**Table 3.** Multivariate analysis for respiratory complications.

| Factors | P value |
| --- | --- |
| Peld score | .325 |
| Zscore less than 2 | .03 |
| Largeforsize graft | .733 |

### Mortality

3 patients died in malnutrition group and 2 patients in no malnutrition group. There was no statistically significant difference between both the groups in overall mortality rates. (p=0.278).

## Discussion

Pre transplant malnutrition is associated with worse post transplant outcomes. Barshes et al showed that worse pre transplant height/age score were associated with increase postoperative hospital stay and hospital costs. (8) Estela in his review stated that malnutrition and growth failure are considered to be important risk factors for poor outcomes following liver transplantation (9). Shephered et al suggested that declining nutrition status in waiting period of transplant adversely affected outcome (10).

Sullivan et al suggested that in malnourished children pre transplant parenteral nutrition gives post transplant outcome comparable to the non-malnourished children (11)

The body weight of children with liver disease can be modulated by many factors, including ascites so weight/age ratio can be confounded by various factors such as ascites, edema in end stage liver disease. So to avoid this confusion and to accurately define pre transplant malnutrition we defined malnutrition as both heights by age and weight by age −2 [wasted and shunted as per who definition] to accurately measure malnutrition.

We compared post transplant complications and mortality between malnutrition or growth failure group and normal group. Complications like bile leak, stricture, bleeding complications, graft dysfunction, Acute cellular rejection, renal dysfunction, sepsis, hepatic artery thrombosis, portal vein stenosis and thrombosis, out flow obstruction, multi organ dysfunction, abdominal compartment syndrome, re-transplantation, intestinal perforation, prolonged ascites etc. were similar between malnutrition and non malnutrition group. In our study malnourished group had significantly higher rate of pulmonary complications (p=0.003). We included complications like ARDS, post transplant pneumonia and atelectasis in pulmonary or respiratory complications. Figueiredo et all suggested in adult liver transplant preoperative malnutrition is associated with an increased risk of post-operative infections, respiratory complications and a prolonged stay in the ICU (12). Poor nutrition induces muscle weakness and muscle cachexia. Prolonged and severe muscle cachexia involving the respiratory muscles may increase postoperative respiratory complications (13).

We also analyzed factors like age at transplant, sex, timing of previous kasai procedure, no of kasai procedure, PELD score, CTP score, Large for size graft, blood loss during surgery, duration of surgery, type of graft for their association with postoperative pulmonary complications. On univariate analysis PELD score and Z score = −2 were significantly associated with respiratory complication. As mentioned above we added Large for size graft in multivariate analysis, as it was significantly different between two groups. On multivariate analysis Z score = −2 independently predicted post transplant respiratory complications. (P value =0.022).

Neto et al in their study of 430 pediatric liver transplants suggested that height by age z score of −2 was associated with poor patient and graft survival (14). Derusso et al showed that after pretransplant weight deficit is associated with late patient mortality (**>**1 year) (15) and soltys et al showed that the baseline z score is related to late growth retardation (16).

This study shows that preoperative optimization of nutritional status may help in getting better out come post transplant in pediatric liver transplant for biliary atresia.

However this study has several limitations, as it is a retrospective study so inherent bias with any retrospective study also applies to this study. Pulmonary complications depend on many aspects like fluid overload; anesthetic management etc., accurate collections of these data on retrospective study are always not possible.

## Conclusion

Pre transplant malnutrition and growth failure is associated with worsened outcome in biliary atresia patients undergoing living donor liver transplantation. Pre transplant malnutrition independently predicts post transplant pulmonary complications. Pre operative optimization of nutrition status might improve outcomes in biliary atresia patients undergoing living donor liver transplantation.

## Data Availability

data can be made available on demand

